# The clinical and genetic spectrum of paediatric speech and language disorders in 52,143 individuals

**DOI:** 10.1101/2024.04.23.24306192

**Authors:** Jan Magielski, Sarah M. Ruggiero, Julie Xian, Shridhar Parthasarathy, Peter Galer, Shiva Ganesan, Amanda Back, Jillian McKee, Ian McSalley, Alexander K. Gonzalez, Angela Morgan, Joseph Donaher, Ingo Helbig

## Abstract

Speech and language disorders are known to have a substantial genetic contribution. Although frequently examined as components of other conditions, research on the genetic basis of linguistic differences as separate phenotypic subgroups has been limited so far.

Here, we performed an in-depth characterization of speech and language disorders in 52,143 individuals, reconstructing clinical histories using a large-scale data mining approach of the Electronic Medical Records (EMR) from an entire large paediatric healthcare network.

The reported frequency of these disorders was the highest between 2 and 5 years old and spanned a spectrum of twenty-six broad speech and language diagnoses. We used Natural Language Processing to assess to which degree clinical diagnosis in full-text notes were reflected in ICD-10 diagnosis codes. We found that aphasia and speech apraxia could be easily retrieved through ICD-10 diagnosis codes, while stuttering as a speech phenotype was only coded in 12% of individuals through appropriate ICD-10 codes. We found significant comorbidity of speech and language disorders in neurodevelopmental conditions (30.31%) and to a lesser degree with epilepsies (6.07%) and movement disorders (2.05%). The most common genetic disorders retrievable in our EMR analysis were *STXBP1* (*n*=21), *PTEN* (*n*=20), and *CACNA1A* (*n*=18). When assessing associations of genetic diagnoses with specific linguistic phenotypes, we observed associations of *STXBP1* and aphasia (*P*=8.57 x 10^-7^, CI=18.62-130.39) and *MYO7A* with speech and language development delay due to hearing loss (*P*=1.24 x 10^-5^, CI=17.46-Inf). Finally, in a sub-cohort of 726 individuals with whole exome sequencing data, we identified an enrichment of rare variants in synaptic protein and neuronal receptor pathways and associations of *UQCRC1* with expressive aphasia and *WASHC4* with abnormality of speech or vocalization.

In summary, our study outlines the landscape of paediatric speech and language disorders, confirming the phenotypic complexity of linguistic traits and novel genotype-phenotype associations. Subgroups of paediatric speech and language disorders differ significantly with respect to the composition of monogenic aetiologies.

## Introduction

Speech and language differences are common clinical features associated with neurodevelopmental disorders. Differences in the neurological basis of communication have been characterized in individuals with specific neurodevelopmental conditions, including rare genetic disorders such as *GRIN2A*-related disorders, *FOXP2*-related disorders, and *STXBP1*- related disorders.^1–4^ There has been promising recent work that has identified novel monogenic and polygenic aetiologies of speech disorders.^5–7^ However, there is still much of the genetic landscape to be elucidated. Accordingly, this represents a major gap in our understanding of speech and language disorders given their presumed genetic component.^8,9^

With the widespread use of Electronic Medical Records (EMR), it becomes possible to systematically study conditions that have not yet received significant attention previously. In addition to making it possible to analyse data on these conditions at scale, EMR allows for the analysis of clinical data over time. For speech disorders in children, this longitudinal component is particularly important given the dynamic nature of neurodevelopment in childhood and adolescence. Hence, as there remains a need to characterize the full clinical spectrum of individuals with communication disorders and the underlying genetic aetiology that impacts differences in speech and language development, EMR-based approaches offer unprecedented opportunities to conduct targeted deep phenotypic analyses at scale.^10,11^

Paediatric speech disorders that have been investigated in the context of their genetic aetiologies include (1) childhood apraxia of speech, (2) childhood dysarthria, and (3) stuttering.^5,8^ *FOXP2* was the first gene discovered to be associated with specific speech impairments, namely speech apraxia and dysarthria.^12–14^ Since this characterization, a variety of genetic aetiologies have been suggested to be associated with neurobiological disruptions of speech and language, but these studies often lack the statistical support that is now available through our increased understanding of population genetics and the development of human genome databases.

Here, we utilized the wealth of information captured in the EMR at a large paediatric specialty care network—including robust primary care, speech-language pathology, developmental, and neurology departments and clinics—to retrieve and reconstruct the longitudinal clinical histories of 52,143 individuals with documented speech and language disorders. A subset of analysis was done on targeted epilepsy and neurogenetics cohorts. We tracked clinical features over time across cohorts and developed a framework for the prediction and identification of clinical subgroups with shared trajectories, allowing us to identify previously unrecognized clinical patterns and to build a more comprehensive understanding of the prevalence and landscape of communication disorders.

## Materials and methods

### Study inclusion and setting

The study was performed at the Children’s Hospital of Philadelphia through the analysis of EMR. We selected a group of the relevant International Classification of Diseases, Tenth Revision (ICD-10) codes: F01-F99, G00-G99, R25-R29, R47-R49, R62, Z13, Z14-Z15, Z81, Z84, I69 to define a broad neurological cohort.^15^ Subsequently, we compiled a list of ICD-10 codes describing speech phenotype-related diagnoses (F80, R47-R49) to delineate our speech cohort (**Supplementary Table 1**). Within this group, we then analysed ICD-10 codes that co-occurred with speech ICD-10 codes to assess their comorbidity with other neurological diagnoses: neurodevelopmental disorders (F84, F88, F89), epilepsy (G40), and movement disorders (G20-G26). We were able to extract the genetic diagnoses individuals from the broad neurological cohort from the dedicated ICD-10 code (Z15.89).

With regards to speech motor disorders, we particularly focused on three including (1) speech apraxia, (2) speech dysarthria, and (3) stuttering. These conditions all fall under the subdivision of motor/neurological speech disorders, as per the classification of the American Speech-Language-Hearing Association.^16^ Speech apraxia is characterized by a difficulty with producing sounds needed for correct pronunciation and an inability to appropriately use prosody in the absence of muscle weakness.^6^ This disorder, however, can co-occur with dysarthria, a condition associated with neuromuscular issues, like abnormal tone, spasticity, or ataxia, which makes the production of comprehensive speech more difficult.^6,17^ Lastly, stuttering is a block in speech fluency which includes features such as repetitions, prolongations, and blocks during fluent speech.^18^

### Patient cohorts and data extraction

In the sub-cohort comprised of individuals from the Pediatric Epilepsy Learning Health System (PELHS) and Epilepsy Genetics Research Project (EGRP), we analysed charts from all encounters; PELHS containing de-identified EMR data of individuals that were seen in our healthcare network and received an epilepsy diagnosis, and EGRP with paediatric patients who are known or believed to have a genetic epilepsy or neurodevelopmental disorder. We extracted phenotypic data using Clinical Text and Knowledge Extraction System, a natural language processing tool, that were then mapped onto the Human Phenotype Ontology (HPO) terms.^19^ This was performed independently from the ICD-10 extraction. By using a well-established controlled dictionary of HPO, we were able to not only record phenotypic information in a standardized computable manner, but also harmonize our dataset, as employed by our group in the past.^20,21^ For example, if a chart of a given individual contained information about stuttering (HP:0025268), this framework enabled us to reason that the individual also had “Abnormality of speech or vocalization” (HP:0002167). Such methods allowed us to simultaneously capture broad and granular phenotypic information—ensuring a thorough phenotypic picture for each individual.

### EMR Visibility Index

Further, we developed a novel EMR Visibility Index by comparing the frequency of clinical speech diagnoses based on the ICD-10 codes against the frequency of speech disorders mentioned in the full-text clinical notes that were mapped onto HPO terms. We developed this novel measure in response to the need of capturing as much EMR data as possible, while accounting for the ‘blind spots’ of this method by identifying clinical groups that tend to be under-characterized because of low visibility in the medical charts. This disparity is particularly important in rare disease communities, who frequently advocate for the creation of new ICD-10 codes for rare conditions so that providers and researchers may reliably track individuals with a given disorders across institutions.^22^ The EMR Visibility Index allowed us to identify the extent of visibility for neurological disorders and speech impairment diagnostic codes, and how their visibility changes depending on the depth of phenotypic analysis. To that end, in the PELHS sub-cohort, we counted the individuals with seizures, speech apraxia, aphasia, autism, intellectual disability, attention deficit hyperactivity disorder (ADHD), and stuttering who had their diagnosis recorded in ICD-10 codes and divided that number by the number of individuals that had the diagnosis coded in their medical charts in HPO. This proportion gave us the EMR Visibility Index.

### Data abstraction and genomic analysis

The documentation and analysis of neurological features associated with speech and language disorders was facilitated through clinical data captured in EMR. Data collected included clinical diagnoses (ICD-10 codes), phenotypic features, neurodevelopmental histories, and genetic findings and diagnoses. Clinical features were mapped across the age-span for all individuals. Further, in 726 individuals from the EGRP sub-cohort—among which 541 individuals had a speech phenotype—we analysed raw exome data from whole-exome sequencing. The raw data alignment, calibration, annotation, and analysis was performed according to the procedure described by our group previously, with the additional step of the Variant Effect Predictor.^23^ Variants that had < 0.005 Genome Aggregation Database (gnomAD) frequency were classified into three groups: class 1— protein-truncating variants (PTVs) with a probability of loss-of-function (pLI) score > 0.95, class 2— Combined Annotation Dependent Depletion (CADD) score > 20 and Residual Variation Intolerance Score (RVIS) < 65, and class 3—PTVs and missense combined. For individuals who had an established genetic diagnosis, only the gene from the genetic diagnosis was used in the analyses to avoid obtaining spurious relationships between other variants that these individuals had with specific speech phenotypes for which their genetic diagnosis would account. Using the Online Mendelian Inheritance in Man (OMIM) database, we identified genes with and without known phenotypic associations.^24^ Variants without known phenotypic associations were then further analysed by evaluating the frequency in gnomAD population database, and through Integrative Genomics Viewer (IGV) to assess reliable alignment of the exome sequencing reads.^25,26^ Further, variants were filtered based on the RVIS to help in prioritizing functional relevance.^27^ Lastly, we leveraged the Database for Annotation, Visualization and Integrated Discovery (DAVID) 2021 bioinformatics resources to better understand possible functional and physiological correlates within our findings.^28^ Reactome Pathway annotation results were employed and, following the DAVID guidelines, looked at the fold enrichment level of above 1.5.^29,30^ If this condition was met, we further analysed whether a nominal *P*-value is significant (< 0.05) for a given association within the DAVID analysis and whether at least 5 genes are present in a given pathway. We subsequently explored the associations of such genes to speech phenotypes in our genotype-phenotype analysis.

### Statistical analysis

All statistical analyses were conducted using the R Statistical Framework.^31^ Statistical testing of associations of Fisher’s exact test is reported with correction for multiple comparisons using False Discovery Rate (FDR) of 5%. If statistical significance was not achieved following correction for multiple comparisons, results were described using their respective odds ratios (OR) with 95% confidence intervals (CI) provided. To assess the similarity of clinical sub-groups within the speech cohort as well as those with and without a genetic diagnosis in the speech and language cohort, Welch two sample t-test was performed. Apart from the FDR of 5%, in the analysis of associations between specific variants and speech and language phenotypes, only variants that were seen in at least two individuals were designated as significant.

## Results

### Speech and language disorders span a wide range of clinical diagnoses

In a broad paediatric cohort of 5,519,989 encounters from 265,926 individuals with a neurological diagnosis, based on twenty-six ICD-10 codes, we identified 1,671,257 encounters across 52,143 individuals with speech and language disorders, spanning a total of 203,150 patient-years (**Fig. 1**). Among these individuals, we found that the most common speech-related ICD-10 diagnoses were mixed receptive-expressive language disorder (F80.2; *n*=27,057 individuals), developmental disorder of speech and language, unspecified (F80.9; *n*=17,579 individuals), expressive language disorder (F80.1; *n*=9,865 individuals). These diagnoses were followed by functional speech sound disorders: phonological disorder (F80.0; *n*=6,060 individuals) and dysphonia (R49.0; *n*=3,184 individuals). The five most common speech disorders accounted for over four-fifths (81.53%) of all speech diagnoses in the cohort. For motor speech disorders with a presumed genetic basis, speech apraxia (R48.2) was seen in 1,099 individuals, stuttering (F80.81) in 1,684 individuals, and dysarthria in 1,056 individuals (R47.1); ICD-10 codes for these disorders represented 4.91% of all speech and language-specific phenotypes. We further observed that among speech and language phenotypes, speech apraxia and aphasia had the highest EMR Visibility Indices (0.74, 0.52), while stuttering had the lowest EMR Visibility Index: 0.12 (**Fig. 2B**).

**Figure 1.**
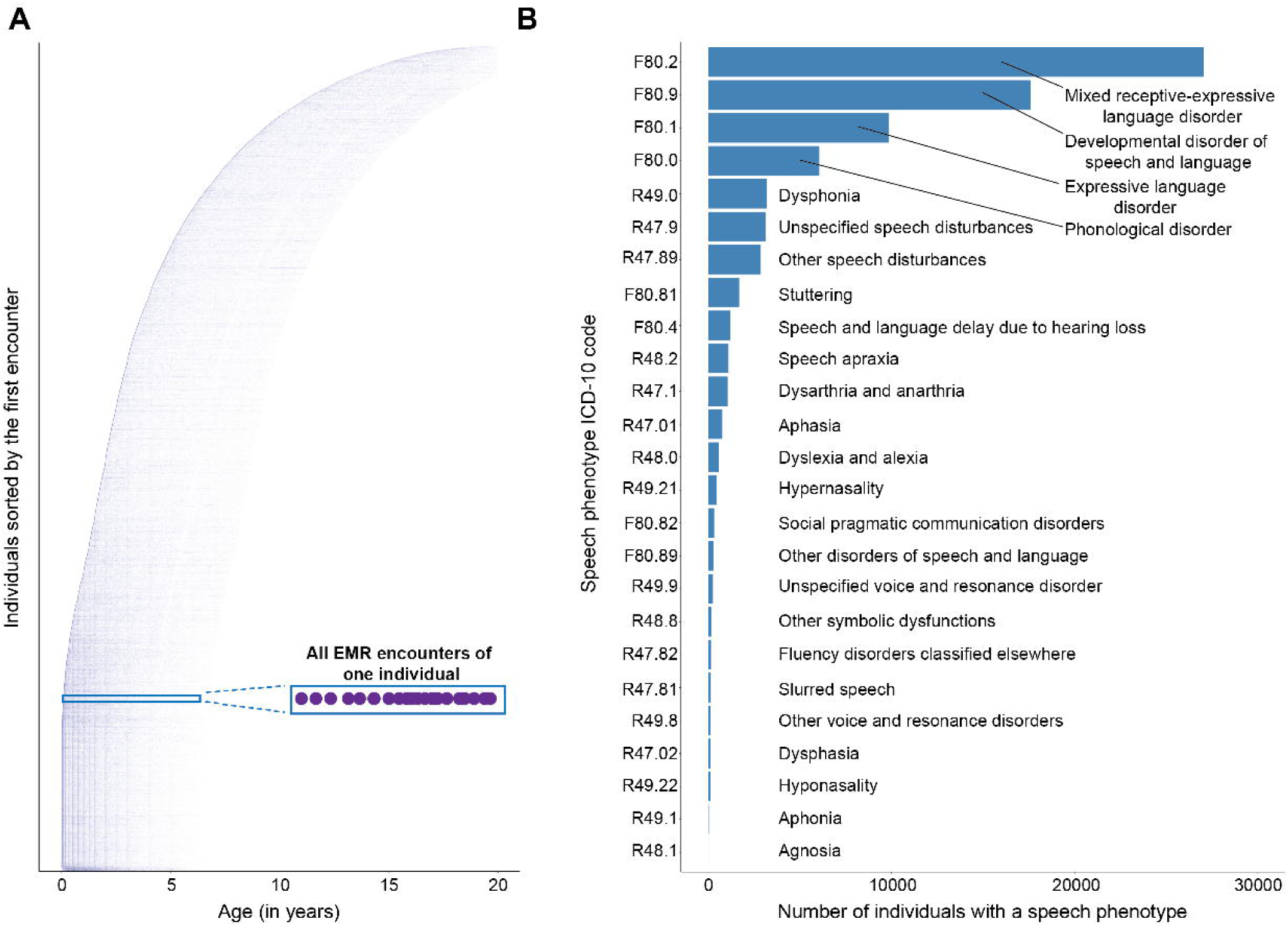
Overview of the speech cohort. **(A)** 1,671,257 encounters across 52,143 individuals with speech disorders, including data from a total of 203,150 patient years. **(B)** Distribution of speech diagnoses.

**Figure 2.**
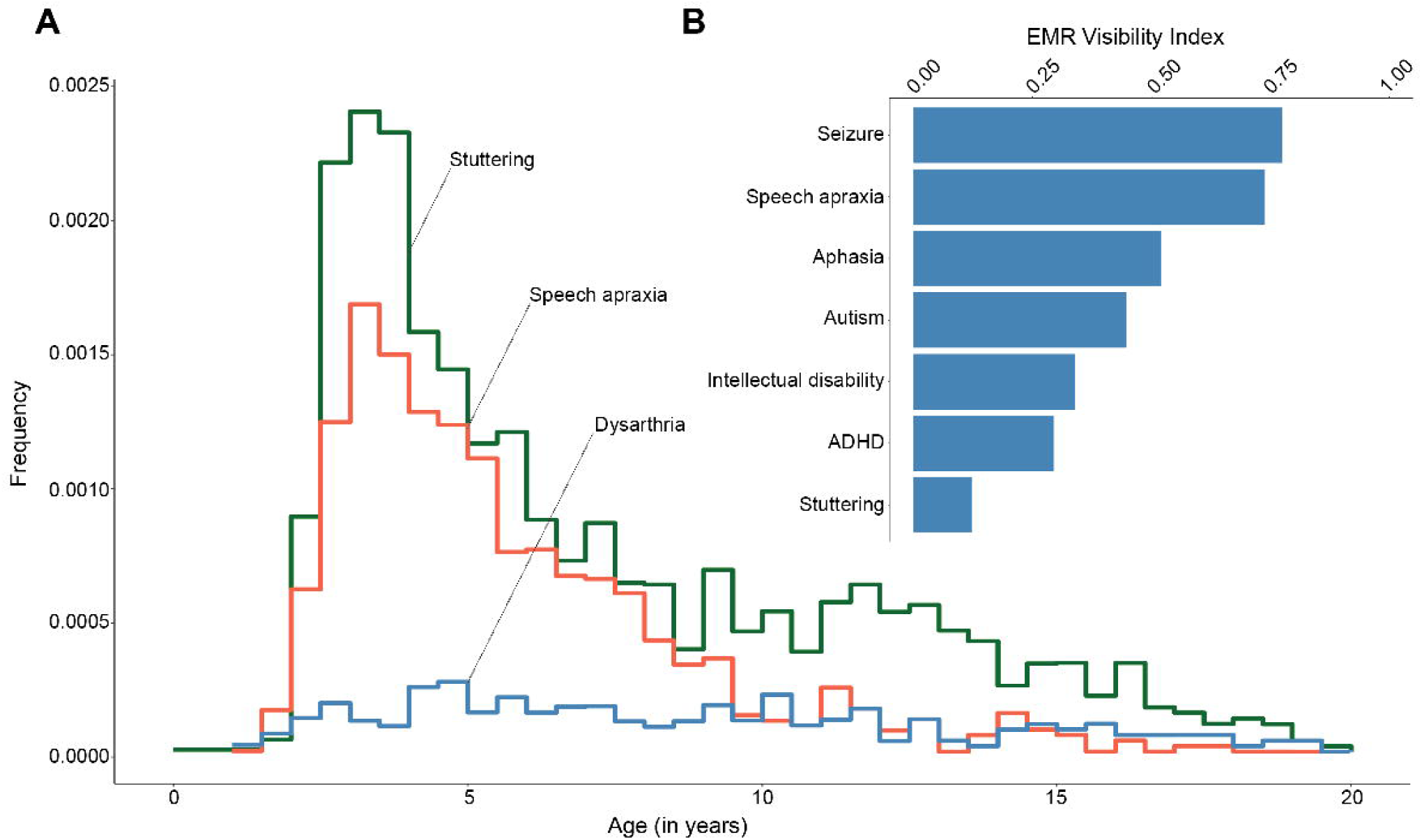
Frequency of specific speech phenotype diagnoses. **(A)** The frequency of stuttering (green), speech apraxia (orange), and dysarthria (blue) diagnoses in 265,926 patients with a neurological diagnosis. **(B)** EMR Visibility Index plot.

### The landscape of speech and language disorders is characterized by age-related phenotypes

We observed that speech phenotype-related diagnoses were most prevalent in the second year of life, with the majority of speech and language diagnoses made between ages 2 and 5, and the highest frequencies seen at two years of age (0.173, *n*=10,938 individuals), one year of age (0.134, *n*=7,924 individuals), and three years of age (0.109, *n*=6,767 individuals). After three years of age, the frequency of speech phenotype-related diagnoses dropped dramatically and was found in less than 10% of all individuals. Within the sub-cohorts of individuals who experience stuttering, speech apraxia, and dysarthria, we observed that the highest frequency still occurs within the 2-5 years old window but slightly later than in the case of paediatric speech and language phenotypes at large (**Fig. 2A**). The frequency of individuals diagnosed with stuttering (frequency in the broad neurological cohort = 0.0141) or dysarthria (frequency in the broad neurological cohort = 3.95 x 10^-4^) reached its peak at 3-4 years of age. Apraxia diagnoses reached its peak at 2-3 years of age (frequency in the broad neurological cohort = 2.09 x 10^-3^). We found that 90% of individuals with a speech abnormality received their first speech and/or language diagnosis by 10.77 years of age.

### Speech disorders overlap with neurodevelopmental disorders, epilepsies, and movement disorders

Examining ICD-10 code diagnoses co-occurring with speech and language phenotypes, we assessed the landscape of speech and language disorders relative to other neurological and psychiatric diseases. We observed the strongest overlap with neurodevelopmental diagnoses: among 52,143 individuals with a speech diagnosis, 15,806 (30.31%; *P* < 2.2 x 10^-16^, OR 6.57, CI 6.40-6.74) also had a neurodevelopmental diagnosis. In our speech cohort, the most frequent co-existing developmental disorders were autism (F84.0: *n*=11,940) and other disorders of psychological development (F88: *n* = 7,239). Epilepsy was found to be the second-most substantial comorbidity (*n*=3,080, 6.07%; *P* = 0.0132, OR 1.05, CI 1.01-1.10) among the broad neurological disorders, with the following most prevalent phenotypes: G40.909: Epilepsy, unspecified (*n*=1,587), G40.209: Focal epilepsy with complex focal seizures (*n*=896), G40.109: Focal epilepsy with simple partial seizures (*n*=847). Lastly, we investigated the overlap between speech and movement disorders, which represented 2.05% of comorbidities (*n*=1,070; *P* = 0.443, OR 0.97, CI 0.91-1.04). Among these, G24.9: Dystonia, unspecified (*n*=290) was the most frequent, followed by G25.3: Myoclonus (*n*=214) and G25.81: Restless legs syndrome (*n*=172; **Fig. 3**).

**Figure 3.**
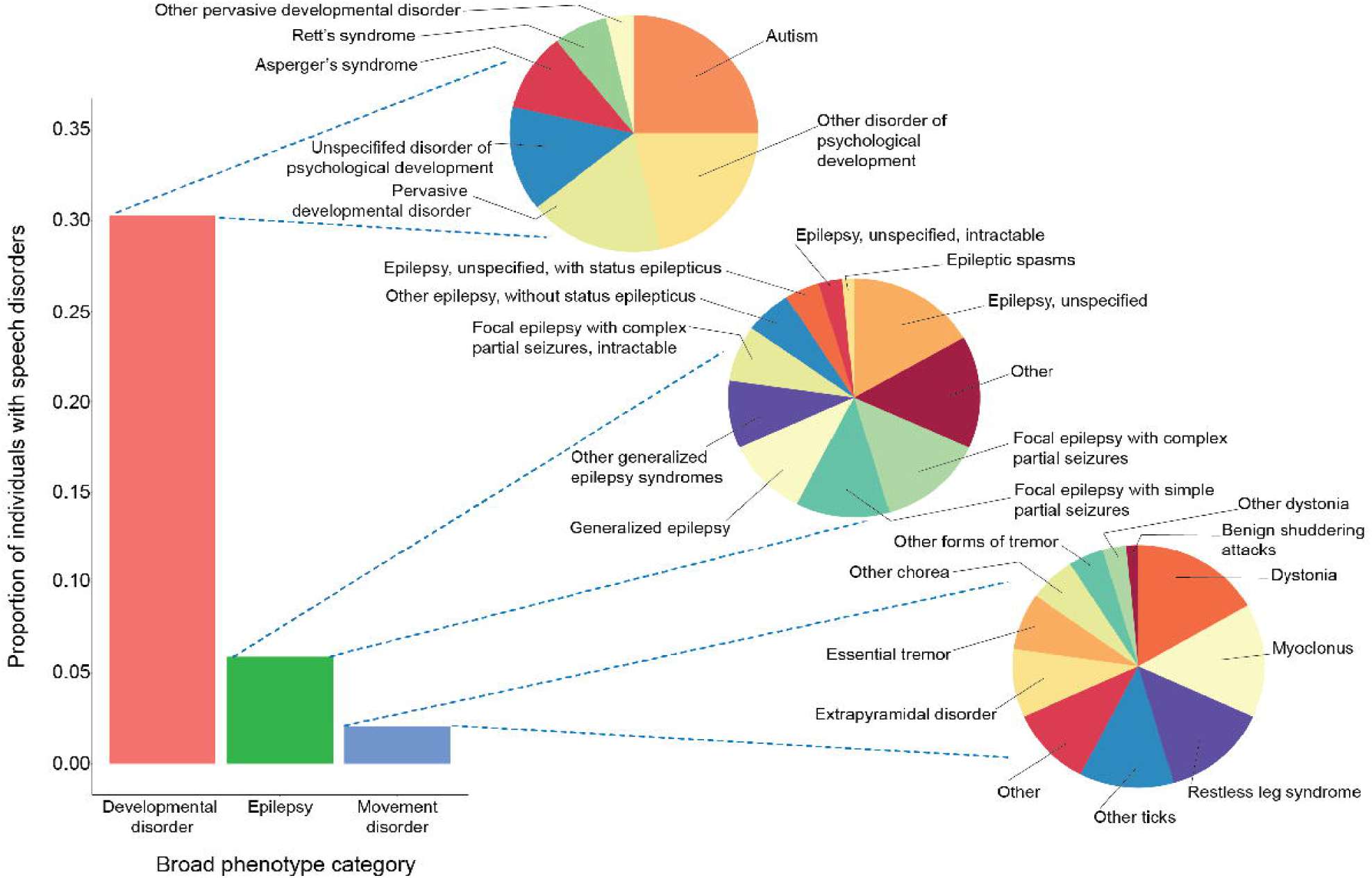
Diagnoses comorbid with speech phenotypes. Broad phenotypic categories co-occurring with speech diagnoses and most common developmental disorder, epilepsy, and movement disorder diagnoses in the speech cohort.

Next, we analysed how these broader co-existing phenotypes related to the age at which the first speech and language diagnoses were made. In the subgroup with comorbid speech and epilepsy diagnoses, 90% of individuals had a speech diagnosis documented by 14.6 years (mean age of diagnosis = 6.80), while for those in the speech and language cohort without an epilepsy diagnosis, that age was 10 years (mean age of diagnosis = 4.44); this difference in the diagnosis age distributions was also captured in the Welch two sample t-test (*P*<2.2 x 10^-16^). Conversely, in the speech-neurodevelopmental sub-cohort including individuals with co-occurring speech and neurodevelopmental disorders, 90% of individuals received their speech diagnosis at 10.2 years (mean age of diagnosis = 4.62), in comparison to 10.7 years for the individuals presenting with a speech phenotype, but without a neurodevelopmental disorder (mean age of diagnosis = 4.56). The difference between the mean age of diagnosis was not significant between the two groups (*P* = 0.096). The data might be limited by the under-documentation of speech phenotypes or the lack of availability of the entirety of the EMR data through one healthcare network system.

### Specific speech and language phenotypes are associated with various genetic aetiologies

We next investigated the landscape of genetic diagnoses in our speech cohort. We found 273 unique genetic diagnoses found in at least one individual, and a total of 607 individuals (1.16%) with a genetic diagnosis. Analysis of cumulative onset of age at which speech diagnoses were first reflected in the EMR demonstrated that 90% of individuals with both a speech/language and genetic diagnosis had documentation of both diagnoses by 12.0 years (mean age = 5.23). The accrual of speech diagnosis occurred slightly later compared to individuals without a genetic diagnosis (90% at 10.5 years, mean age = 4.57, **Fig. 4**); the distribution of speech diagnosis age was significantly different between the two groups, as evidenced by the Welch two sample t-test (*P* = 0.0002). The most common genetic diagnoses included *STXBP1* (*n*=21), *PTEN* (*n*=20), *CACNA1A* (*n*=18), *SCN2A* (*n*=14), and *SYNGAP1* (*n*=11). We next explored more granular relationships between specific speech and language disorder types and genetic diagnoses. After correcting for multiple testing, the following relationships were significant: *STXBP1* with aphasia (*P* = 8.57 x 10^-12^, OR 50.23, CI 18.62-130.39) as well as *MYO7A* with other developmental disorders of speech and language (*P* = 1.24 x 10^-5^, OR Inf, CI 17.46-Inf). The nominally significant relationships with the highest level of significance included *GRIN2A* with speech apraxia (*P* = 3.3 x 10^-4^, OR 34.06, CI 4.98-201.11), *MECP2* with other developmental disorders of speech and language (*P* = 9.81 x 10^-4^, OR 54.02, IC 5.45-284.24), and *POLG* (*P* = 0.0013, OR 65.87, CI 4.77-898.38) with aphasia (**Fig. 5**, **Table 1**).

**Figure 4.**
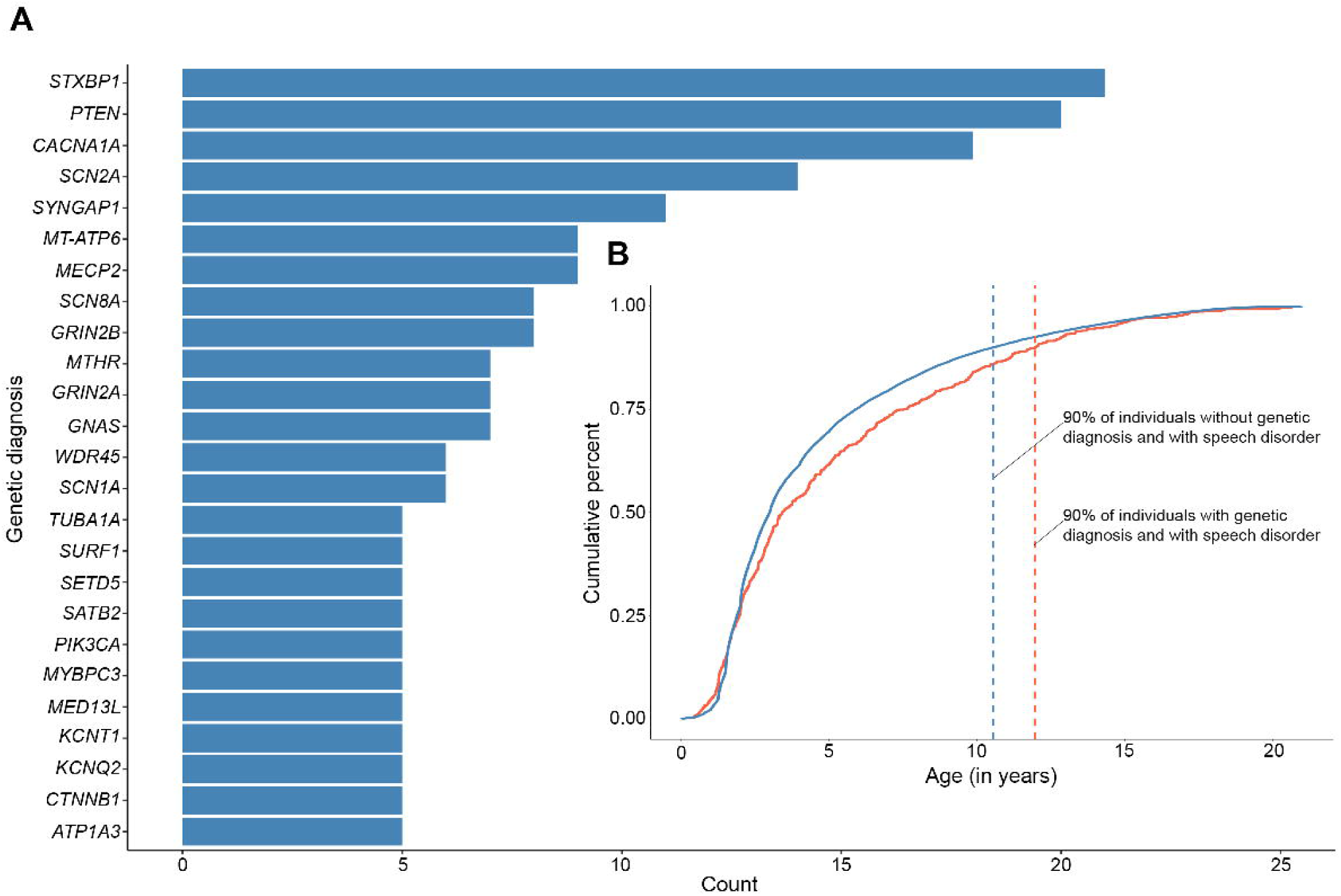
Genetic diagnoses in the speech cohort. **(A)** Distribution of the genetic diagnoses with n>=5 in the speech cohort (left) and cumulative onset of speech diagnosis in individuals with (orange) and without (blue) a genetic diagnosis (in-set).

**Figure 5.**
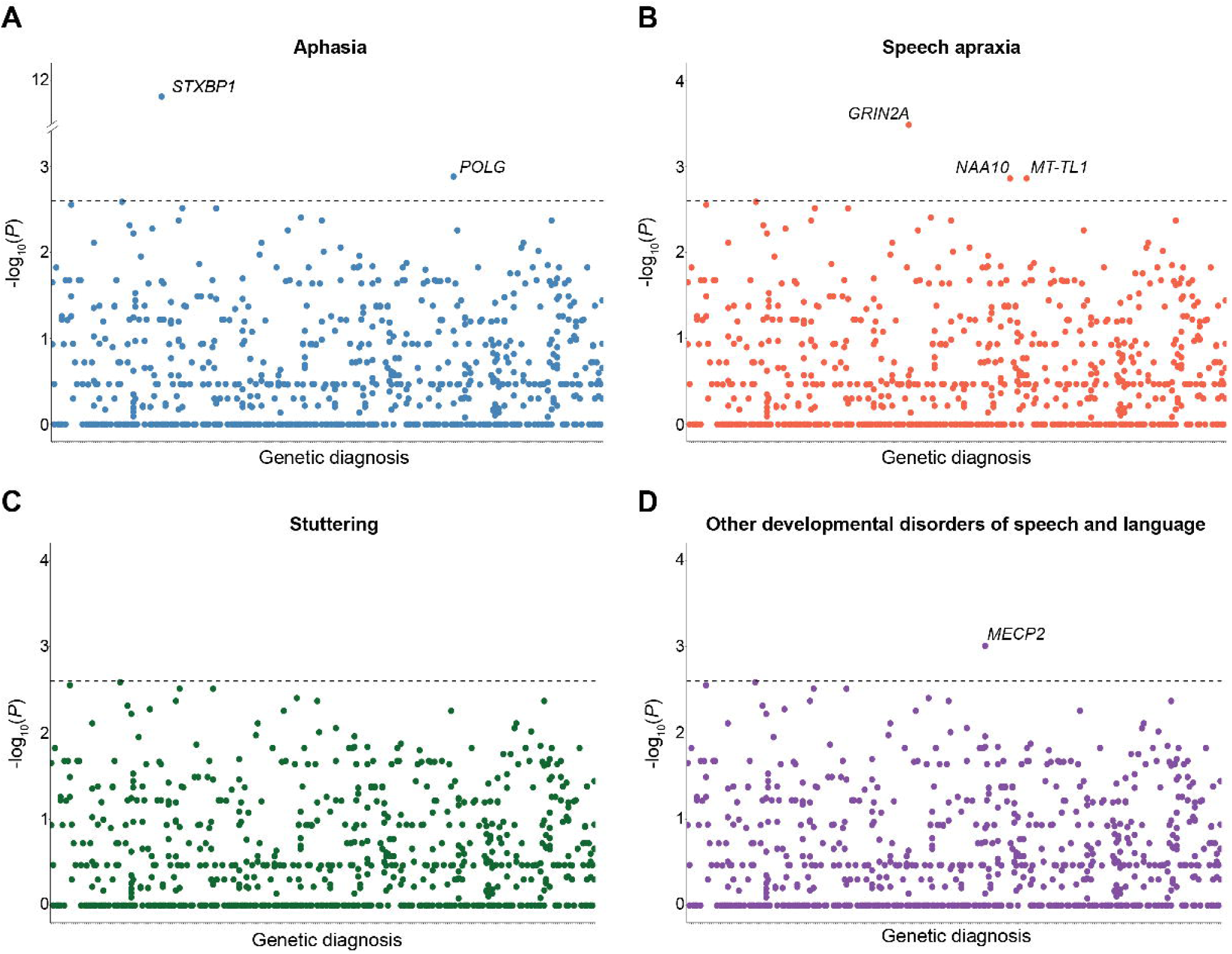
Associations between clinical genetic diagnoses and speech phenotypes. **(A)** aphasia, **(B)** speech apraxia, **(C)** stuttering, and **(D)** other developmental disorders of speech and language. Labels are assigned to the diagnoses that maintained significance after FDR of 10% after exploratory analysis shown by the dashed line.

**Table 1.** Associations between genetic diagnoses and speech phenotypes.

| **Aphasia (R47.01)** |  |  |  |  |  |
| --- | --- | --- | --- | --- | --- |
| Genetic diagnosis | Individuals | P-value | OR | 95% CI | Frequency |
| *STXBP1* | 9 | 8.57 x 10<sup>-12</sup>* | 50.23 | 18.62-130.39 | 0.43 |
| *POLG* | 2 | 0.0013 | 65.87 | 4.77-898.38 | 0.5 |
| *CACNA1C* | 1 | 0.0297 | 65.79 | 0.84-4911.20 | 0.5 |
| *APC* | 1 | 0.0443 | 32.86 | 0.56-630.79 | 0.33 |
| *TUBA1A* | 1 | 0.0727 | 16.46 | 0.33-166.81 | 0.2 |
| **Speech apraxia (R48.2)** |  |  |  |  |  |
| *GRIN2A* | 3 | 3.30 x 10<sup>-4</sup> | 34.06 | 4.98-201.11 | 0.43 |
| *NAA10* | 2 | 0.0014 | 90.60 | 4.71-5110.56 | 0.67 |
| *MT-TL1* | 2 | 0.0014 | 90.52 | 4.71-5106.15 | 0.67 |
| *CACNA1C* | 1 | 0.0428 | 45.21 | 0.576-3430.25 | 0.5 |
| *GABRB3* | 1 | 0.0428 | 45.21 | 0.576-3430.25 | 0.5 |
| **Dysarthria and anarthria (R47.1)** |  |  |  |  |  |
| *NKX2-1* | 2 | 0.0013 | 92.88 | 4.83-5232.37 | 0.67 |
| *NUBPL* | 2 | 0.0013 | 92.88 | 4.83-5232.37 | 0.67 |
| *KCNQ2* | 2 | 0.0043 | 30.93 | 2.58-270.02 | 0.4 |
| *CTNNB1* | 2 | 0.0043 | 30.93 | 2.58-270.02 | 0.4 |
| *SURF1* | 2 | 0.0043 | 30.93 | 2.58-270.02 | 0.4 |
| **Speech and language development delay due to hearing loss (F80.4)** |  |  |  |  |  |
| *MYO7A* | 3 | 1.24 x 10<sup>-5</sup>* | Inf | 17.46-Inf | 1 |
| *GJB2* | 2 | 0.0016 | 84.97 | 4.42-4807.29 | 0.67 |
| *KCNQ1* | 2 | 0.0016 | 84.89 | 4.41-4803.38 | 0.67 |
| **Other developmental disorders of speech and language (F80.89)** |  |  |  |  |  |
| *MECP2* | 2 | 9.81 x 10<sup>-4</sup> | 54.02 | 5.45-284.24 | 0.22 |
| *GLI3* | 1 | 0.0106 | 187.91 | 2.38-12642.68 | 0.5 |
| *PACS1* | 1 | 0.0159 | 93.49 | 1.58-1817.74 | 0.33 |
| *DYRK1A* | 1 | 0.0211 | 62.60 | 1.19- 795.86 | 0.25 |

\*If significant after the FDR correction for multiple testing.

### Exome sequencing analysis further shows there is an underlying genetic component to speech and language disorders

As expected, analysis of exome sequencing data in 726 individuals revealed a variety of rare variants. In total, we found 212 PTVs (class 1), 6,355 variants with CADD score > 20 and RVIS < 65 (class 2), and 15,181 variants in the combined PTV-missense group (class 3); 95 (13.09%) individuals had a clinically verified genetic diagnosis. We observed that variants in the following genes were significantly associated with speech phenotypes after the correction for multiple testing: *UQCRC1*-expressive aphasia and *WASHC4*-abnormality of speech or vocalization (**Fig. 6**, **Table 2**). PTVs in the following genes showed nominally significant relationships with speech and/or language phenotypes: *SMARCE1*-aphasia (*P* = 0.0103, OR 45.00, CI 1.67-Inf), *RERE*-receptive language delay (*P* = 0.0166, OR 18.00, CI 1.26-250.85), *SMARCE1*-dysarthria (*P* = 0.0228, OR 28.41, CI 1.06-Inf), *MAZ*-stuttering (*P* = 0.0362, OR 11.45, CI 0.80-159.30), and *PDPK1*-language impairment (*P* = 0.0418, OR 4.97, CI 0.83-34.20; **Table 2**).

**Figure 6.**
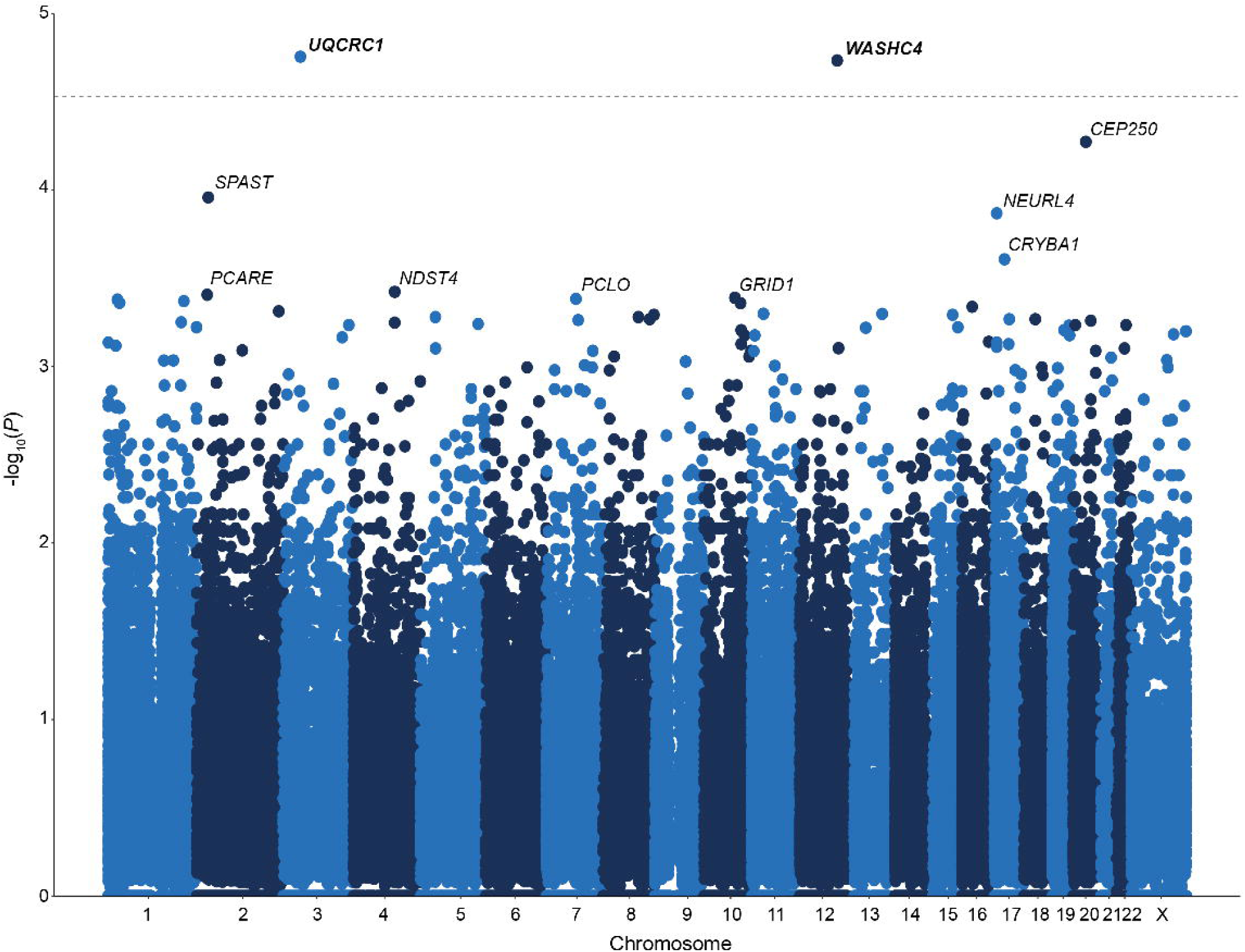
Variants present in the speech cohort. Distribution of all variants in the speech cohort. The dashed line represents the post-FDR threshold of significance, and the names of the genes with significant associations are bolded.

**Table 2.**
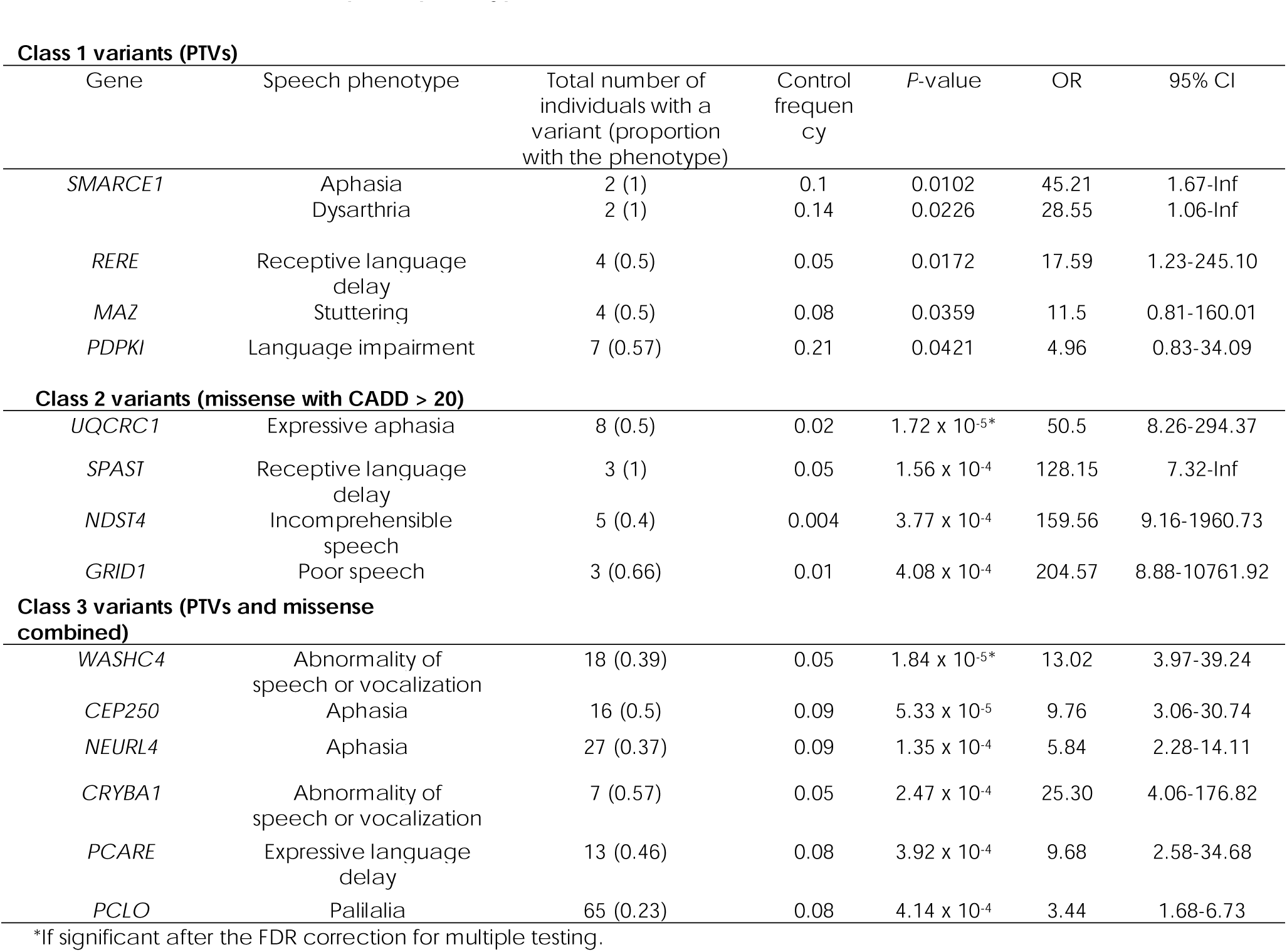
Associations between speech phenotypes and rare variants. Class 1 variants (PTVs)

### Genes contributory to speech disorders cluster in neurologically relevant pathways

We broadly entered all nominally significant class 2 variants (*n*=781) from our cohort— optimal number between 100 and 2,000 per Huang et al.—to DAVID that would allow for a meaningful integrative analysis and a reliable assessment of enrichment relative to background.^29^ From this analysis, we found that genes involved in the synaptic adhesion-like pathway were most enriched (fold enrichment = 5.3, *P* = 0.013); these included *GRIN2A*, *GRIN2B*, *FLOT2*, *PTPRD*, and *PTPRF*. *GRIN2A* and *GRIN2B* were enriched in several other pathways: transcriptional regulation by MECP2 (fold enrichment = 3.6, *P* = 0.0017), protein-protein interaction at synapses (fold enrichment = 2.8, *P* = 0.005), and neuronal system (fold enrichment = 1.5, *P* = 0.03). Other pathways that emerged as significant in terms of their number of genes, *P*-value threshold, and fold enrichment spanned the three main physiological processes: sound processing, cellular structure, and cellular interactions (**Supplementary Table 2**).

## Discussion

In this study, we conducted a comprehensive analysis of the landscape of paediatric speech and language disorders, leveraging clinical information captured from routine care within EMR of 52,143 individuals across 203,150 years of patient data at a major US paediatric academic hospital. Overall, through this high-throughput EMR genomics approach, we confirmed the knowledge established previously by traditional phenotyping studies of smaller sample size, while expanding their findings. This approach allowed us to make three crucial observations. First, we found substantial heterogeneity of speech diagnoses, with mixed receptive-expressive language disorder and developmental disorder of speech and language being the most frequent diagnoses. Second, speech and language disorders have considerable overlap with neurodevelopmental disorders, movement disorders, and epilepsy.^13,32^ Lastly, distinct speech phenotypes can be associated with specific genotypic findings and demonstrate genetic overlap with known neurodevelopmental genetic conditions.^2,4,6^

As expected, our analysis of speech diagnoses showed that, even though there were a total of twenty-six ICD-10 codes corresponding to this broad clinical presentation, the broader phenotypic diagnoses were the most frequent. Terms describing mixed receptive-expressive language disorder, developmental disorder of speech and language or expressive language disorder were over 11 times more prevalent than more specific speech disorder ICD-10 codes, like speech apraxia, stuttering, or dysarthria. While the general speech diagnoses are undoubtedly useful in assessing high-level phenotypic associations, parsing out more granular features of speech impairment has proven to be difficult at this level of generalization which accompanies the use of standard EMR. This observation reflects what other researchers in the field have noted about the need for deep speech phenotyping in order to accurately describe this phenotypic landscape, characterize clinical trajectories, and allow for high-yield phenotype-gene associations discoveries.^10^ It is worth noting that speech and language impairment is often considered a feature of neurodevelopmental disorders, rather than an entity of its own, which may be a factor that hinders precise characterization of these conditions. Our analysis supports this observation via the EMR Visibility Index; stuttering, a speech disorder with an elusive genetic underpinning, was least visible when assessing ICD-10 codes in our cohort. Here, only slightly more than one in ten individuals had their stuttering diagnosis reflected in ICD-10 codes. This may account for prior observations that stuttering is a virtually absent diagnosis within large biobanks.^11^ Additionally, it is possible that our data is affected by the fact that many individuals who stutter receive their care through community centers and school-based therapies. In short, this demonstrates that genomic approaches using EMR data may not provide clear insight into a particular phenotype, requiring novel approaches such as phenotype classifiers^33^ or, as in our study, analysis of full-text clinical notes through Natural Language Processing.

While the high-level nature of the speech and language-related ICD-10 codes pose challenges to subsequent analyses, we were able to add additional granularity by analysing longitudinal clinical data through time-stamped progression of clinical trajectories. Hence, we were able to observe that—regardless of how general or specific a given ICD-10 code was—the age period between two and five-years of age was when the frequency of speech diagnoses was the highest, in line with long established epidemiologically confirmed knowledge of child speech and language disorders.^34^

Comorbidity with other conditions is a critical aspect of the phenotypic spectrum of speech and language disorders. We appreciated substantial overlap with neurodevelopmental disorders, which was more than five times as high as that seen with epilepsies or movement disorders. This result is consistent with the general clinical presentation of neurodevelopmental disorders: speech and language impairment is a common domain affected in such conditions.^35^ It is possible that, for this reason, speech and language differences are noticed more frequently in medical records of individuals with neurodevelopmental diagnoses^36^ and is given attention in clinical care in these cases.

The clear relationship between speech and language and neurodevelopmental disorders was also reflected by the spectrum of genetic diagnoses that we observed in our cohort. The genetic diagnoses that we identified here were related to genes known to be contributory in various neurodevelopmental disorders and epileptic encephalopathies; *STXBP1*, *GRIN2A*, *POLG*, and *MECP2*—which is consistent with what was reported in the literature previously.^2,37–39^ Further, genes for which there was a nominally significant association with speech disorders were those contributing to movement disorders: *NKX2-1* is associated with chorea and *NUBPL* with ataxia and dystonia.^40,41^ The last group of genes that showed nominally significant relationship with speech and or language phenotypes were known to be contributory to hearing loss: *GJB2* and *KCNQ1*.^42,43^ The breadth of the genetic diagnoses spectrum illustrates the various dimensions of potential aetiologies of speech impairment, ranging from epileptic encephalopathies to movement disorders and hearing loss, mirroring the findings of our phenotypic-based analysis. Disentangling speech and language phenotype-genotype association warrants further examination; we identified several relationships, but no genes that would be explanatory for speech and language impairments alone were identified in our cohort. It is worth noting that we identified genetic diagnoses with a frequency of occurrence equal to 1 in our cohort (**Supplementary Table 3**). Some of these included genes that are known to be contributing to conditions leading to speech or language impairment, such as *MYO7A* and hearing loss,^44^ as well as other genes that were identified in singular cases in our cohort, but were not reported to be contributory elsewhere. This provides insight to the potential breadth of genes contributing to speech and language phenotypes.

With an increased search radius for both phenotypes, using more granular clinical data extracted from Natural Language Processing of patient notes than clinical diagnoses and genotypes, analysing exome sequencing in lieu of genetic diagnoses, we found more evidence for a genetic basis for speech and language phenotypes. We showed that variants in genes that have, and do not have, an established phenotype were found to contribute to speech and language disorders. Variants in *UQCRC1* have been established to be causative of parkinsonism with polyneuropathy.^45^ Our work further extends the spectrum of the disorders related to deleterious missense variants in this gene, revealing a prominent association with expressive aphasia. Similarly, we identified an association of speech and language phenotypes in individuals with *WASHC4* variants, a gene that had previously been established as a cause of an autosomal recessive developmental disorder. ^46^ In our cohort, the broad phenotype of abnormality of speech or vocalization was found to be associated with heterozygous variants in this gene, which suggests a possibly novel phenotype for *WASHC4* in heterozygotes. In terms of the nominally significant PTV-speech phenotype associations, our analysis revealed that both variants in genes with a known associated development disorder-related phenotype (*SMARCE1*, *RERE*) and without an established clinical presentation (*MAZ*, *PDPK1*) may contribute to speech disorders.^47–49^ *MAZ* encodes a myc-associated zinc finger protein, a transcription factor, which plays an important role in the process of gliogenesis,^50^ while PTVs in *PDPK1* have been previously reported to be associated with autism.^51^

To better understand the biological meaning and functional clustering of variants in genes nominally associated with speech phenotypes, we performed DAVID analyses, which showed that the most enriched pathways constitute central elements of neurologically crucial processes. Firstly, these results confirmed what we established on the level of the ICD10-genetic diagnosis analysis—we observed nominally significant results for *GRIN2A*, *CACNA1C*, and *MYO7A* in both analyses. This exhibits the high quality and sensitivity of the EMR genomics approaches, while highlighting the importance of comprehensive integrative bioinformatic analysis when dealing with rare variants. With these technologies we were able to demonstrate that such rare variants can be grouped into physiologically relevant categories.^29^ This bioinformatic analysis further supported the idea that genetic architecture of speech disorders is related to developmental and hearing loss conditions, as demonstrated by the enriched pathways. Glutamatergic neurotransmission appears to play a particularly prominent role in the genetics of speech impairment.^52^ While it was known before that *GRIN2A* had a characteristic speech and epilepsy phenotype, we determined that *GRIN2B* and *GRM1* are also associated with speech impairment.^2^ This demonstrates a meaningful expansion of the existing knowledge of *GRIN2B-* and *GRM1-*related conditions, which have been previously associated with developmental epileptic encephalopathy and spinocerebellar ataxia, respectively.^53,54^ Though these were absent in the DAVID analysis output, other glutamate receptor genes with both known (*GRIA2*, *GRM7*) and unestablished phenotypes (*GRID1*, *GRIK3*, *GRIN3B*) showed nominally significant associations with speech differences in our exome analyses.^55,56^ This analysis is consistent with what we observed on the phenotypic level through EMR analysis: the nature of speech disorders intersects with that of neurodevelopmental disorders.

To date, this is the first attempt to characterize speech disorders as their own entity and map them using longitudinal EMR data. We demonstrated that they tend to overlap both phenotypically and genetically with developmental, epilepsy, and movement disorders. Novel variants we observed to be associated with speech phenotypes show a possible phenotypic plurality as conditions may have differing clinical characteristics depending on the genetic variation.

Further investigation into the landscape of the genetic architecture of speech disorders is necessary. Prospective studies and genetic testing of individuals affected by such conditions can provide further insights into how variants in specific genes contribute to distinct speech presentations. While we provide a comprehensive perspective on speech phenotypes here, the depth of phenotypic analysis is limited by the EMR-driven methods. Additionally, EMR genomics approaches can be influenced by specific centers of expertise contained within a particular healthcare network. It is possible that some genes causative of epilepsy and neurodevelopmental conditions emerged from our analysis due to a large epilepsy genetics centre at Children’s Hospital of Philadelphia, where children with these diagnoses are seen frequently. Future explorations may pursue phenotyping approaches in a similar computational manner, but in cohorts comprised of individuals with a pre-defined speech disorder (e.g., stuttering, speech apraxia, dysarthria) which would allow for more finite analysis of associations between genetic changes and speech features. Targeted studies as described above are critical for the discovery of novel genotype-phenotype associations, as well as gene discovery, in the realm of speech disorder genetics.

## Data availability

Primary data used in this study are available upon request from the corresponding author.

