## Supplementary material for "The clinical and genetic spectrum of paediatric speech and language disorders in 52,143 individuals"

**Supplementary Table 1 ICD-10 code-based definition of the speech cohort.**

| **ICD-10 code** | **Definition** |
| --- | --- |
| F80.0 | Phonological disorder |
| F80.1 | Expressive language disorder |
| F80.2 | Mixed receptive-expressive disorder |
| F80.4 | Speech and language delay due to hearing loss |
| F80.8 | Other developmental disorders of speech and language |
| F80.81 | Childhood onset fluency disorder (stuttering) |
| F80.82 | Social pragmatic communication disorder |
| F80.89 | Other developmental disorders of speech and language |
| F80.9 | Developmental disorder of speech and language, unspecified |
| R47 | Speech disorders, not elsewhere classified |
| R47.01 | Aphasia |
| R47.02 | Dysphasia |
| R47.1 | Dysarthria and anarthria |
| R47.81 | Slurred speech |
| R47.82 | Fluency disorder in conditions classified elsewhere |
| R47.89 | Other speech disturbances |
| R47.9 | Unspecified speech disturbances |
| R48.2 | Apraxia (speech apraxia) |
| R49.0 | Dysphonia |
| R49.1 | Aphonia |
| R49.8 | Other voice and resonance disorders |
| R49.9 | Unspecified voice and resonance disorder |

**Supplementary Table 2 Reactome pathways enriched in the analysis (with enrichment > 2.5 and *P*-value < 0.05).**

| Reactome pathway (stable identifier) | Genes | Fold enrichment | *P*-value |
| --- | --- | --- | --- |
| Synaptic adhesion-like molecules  (R-HSA-8849932) | *GRIN2A*, *GRIN2B*, *PTPRD*, *PTPRF*, *FLOT2* | 5.3 | 0.013 |
| NCAM1 interactions  (R-HSA-419037) | *CACNB3*, *CACNA1C*, *CACNA1D*, *COL4A2*, *COL5A3*, *COL6A6*, *CNTN2* | 3.7 | 0.011 |
| MET activates PTK2 signalling  (R-HSA-8874081) | *COL1A2*, *COL5A3*, *COL27A1*, *LAMA3*, *LAMA4* | 3.7 | 0.044 |
| Laminin interactions  (R-HSA-3000157) | *COL4A2, HSPG2, ITGA7, LAMA3, LAMA4* | 3.7 | 0.044 |
| Transcriptional regulation by MECP2  (R-HSA-8986944) | *GRIN2A*, *GRIN2B*, *FKBP5*, *RBFOX1*, *AURKB*, *CAMK2D*, *GAD2*, *SGK1*, *TRPC3*, *TNRC6B* | 3.6 | 0.0017 |
| MET promotes cell motility  (R-HSA-8875878) | *COL1A2, COL5A3, COL27A1, DOCK7, LAMA3, LAMA4* | 3.3 | 0.035 |
| ECM proteoglycans  (R-HSA-3000178) | *COMP, COL1A2, COL4A2, COL5A3, COL6A6, HSPG2, ITGA7, LAMA3, LAMA4, TNC* | 2.9 | 0.0068 |
| Sensory processing of sound  (R-HSA-9659379) | *EPS8*, *EPS8L2*, *LHFPL5*, *CDH23*, *CACNA1D*, *EPB4ILI*, *MYO7A*, *MYO15A*, *PCLO*, *KCNQ4* | 2.9 | 0.0068 |
| Assembly of collagen fibrils and other multimeric structures  (R-HSA-2022090) | *COL1A2, COL4A2, COL5A3, COL6A6, COL27A1, LAMA3, LOXL2, LOLXL3* | 2.9 | 0.019 |
| Integrin cell surface interactions  (R-HSA-216083) | *F11R, COMP, COL1A2, COL4A2, COL5A3, COL6A6, HSPG2, ITGA7, KDR, SPP1, TNC* | 2.9 | 0.0046 |
| Protein-protein interactions at synapses  (R-HSA-6794362) | *GRIN2A*, *GRIN2B*, *GRM1*, *PPFIA4*, *EPB41L1*, *EPB41*, *FLOT2*, *IL1RAP*, *PTPRD*, *PTPRF*, *SYT12* | 2.8 | 0.005 |
| Non-integrin membrane-ECM interactions  (R-HSA-3000171) | *COL1A2, COL4A2, COL5A3, HSPG2, LAMA3, LAMA4, TNC* | 2.6 | 0.048 |
| RHOA GTPase cycle  (R-HSA-8980692) | *ARAP2, ARAP3, IQGAP1, IQGAP3, ARHGAP29, ARHGAP32, ARHGEF28, ARHGEF4, SLK, STARD13, ANLN, CCDC115, FAM13A, FLOT2, KALRN, PLEKHG3, VAV1* | 2.6 | 0.00091 |

**Supplementary Table 3 Selection of associations between genetic diagnoses and speech phenotypes with a frequency of one.**

| **Speech phenotype**  **(ICD-10)** | **Genetic diagnosis** |
| --- | --- |
| Aphasia  (R47.01) | *MBD5* (n=1) |
|  | *KCND3* (n=1) |
|  | *GRIN1* (n=1) |
| Speech apraxia  (R48.2) | *PIK3CA* (n=1) |
|  | *SUCLA2* (n=1) |
|  | *CLN6* (n=1) |
| Dysarthria and anarthria  (R47.1) | *AIFM1* (n=1) |
|  | *PIGN* (n=1) |
|  | *NDUFAF6* (n=2) |
| Stuttering  (F80.81) |  |
|  | *TRPV4* (n=1) |
|  | *DICER1* (n=1) |
|  | *DYNC1H1* (n=1) |
|  | *TGFBR1* (n=1) |
| Speech and language development delay due to hearing loss  (F80.4) | *MYO7A* (n=3) |
|  | *GATA3* (n=2) |
|  | *USH2A* (n=2) |
| Other developmental disorders of speech and language  (F80.89) | *CEP290* (n=1) |
